# Systematic Review and Meta-Analysis of the Efficacy and Effectiveness of Pneumococcal Vaccines in Adults

**DOI:** 10.1101/2022.10.06.22280772

**Authors:** Jennifer L Farrar, Lana Childs, Mahamoudou Ouattara, Fahmina Akhter, Amadea Britton, Tamara Pilishvili, Miwako Kobayashi

**Author notes:** Corresponding author: Jennifer L Farrar.

## Abstract

The 13-valent pneumococcal conjugate vaccine (PCV13) and 23-valent pneumococcal polysaccharide vaccine (PPSV23) were previously recommended for adults in the United States. To help inform discussions on recently licensed 15- and 20-valent pneumococcal vaccine use among adults, we conducted a systematic review of PCV13 and PPSV23 efficacy or effectiveness. We conducted a search on PCV13 and PPSV23 efficacy or effectiveness (VE) studies against vaccine type (VT) invasive pneumococcal disease (IPD) and VT-pneumococcal pneumonia in adults. Nineteen studies were included: 13 on VT-IPD (four on PCV13, nine on PPSV23) and eight on VT-pneumococcal pneumonia (three on PCV13, four on PPSV23, one on PCV13 and PPSV23). One randomized-controlled trial (RCT) evaluated PCV13 and observed an efficacy of 75% and 45% against VT-IPD and VT-pneumococcal pneumonia, respectively. No RCTs reported PPSV23 efficacy. PCV13 effectiveness estimates against VT-IPD ranged from 47% to 68%. Pooled PPSV23 effectiveness against VT-IPD was 45% (95% CI: 37%, 51%; I^2^=0%). PCV13 VE estimates against VT-pneumonia ranged from –2 to 46%. Pooled PPSV23 VE against VT-pneumococcal pneumonia was 18% (95% CI: -4%, 35%; I^2^=0%). Evidence suggests PCV13 and PPSV23 are effective against VT-IPD and VT-pneumococcal pneumonia in adults; this was used to inform PCV15 and PCV20 policy decisions.

## Introduction

The bacteria, *Streptococcus pneumoniae*, are responsible for a substantial proportion of disease burden in adults, especially among older adults and those with immunocompromising conditions. Invasive pneumococcal disease (IPD) incidence among U.S. adults aged ≥65 years was 24 per 100,000 during 2018–2019 (ABCs surveillance), and disease caused by serotype 3, a serotype included in vaccines to prevent pneumococcal disease, is the most common vaccine serotype causing IPD in adults [1, 2]. In a 2016 study of U.S. adults hospitalized with radiologically confirmed community-acquired pneumonia, 8.2% (n=520) of adults ≥65 years had *S. pneumoniae* detected by urine antigen; of these, 269 (51.7%) were serotypes included in the 13-valent pneumococcal conjugate vaccine (PCV13) [2].

Until recently, there were two pneumococcal vaccines recommended for use in U.S. adults to prevent disease caused by *S. pneumoniae*. Since 2012, PCV13 has been recommended for use in adults aged ≥19 years with immunocompromising conditions, functional or anatomic asplenia, cerebrospinal fluid leaks, or cochlear implants. The 23-valent pneumococcal polysaccharide vaccine, PPSV23, has been in use since 1984, and, until recently, was recommended for all adults aged ≥65 years and adults aged ≥19 years with immunocompromising conditions, functional or anatomic asplenia, or immunocompetent conditions [3]. While studies have shown that PPSV23 is effective against VTIPD, data on effectiveness of PPSV23 against nonbacteremic pneumococcal pneumonia have been inconsistent [4]. In contrast, a large randomized controlled trial (RCT) in adults aged ≥65 years in the Netherlands showed that PCV13 is effective against VT-IPD and VT-nonbacteremic pneumococcal pneumonia [5]. Data from this study supported the 2014 recommendation for routine PCV13 use for all adults aged ≥65 years in the United States [4]; however, additional years of data showed that there was limited population-level impact of routine PCV13 use in adults of this age group, likely due to the already realized indirect effects from PCV13 use in children. As a result, in 2019, PCV13 was no longer routinely recommended for adults aged ≥65 years without immunocompromising conditions, cerebrospinal fluid leaks, or cochlear implants and instead recommended based on shared clinical decision-making [6].

In 2021, two new pneumococcal conjugate vaccines (PCVs) were licensed for use in the United States: 15-valent PCV (PCV15) and 20-valent PCV (PCV20) [7, 8]. PCV15 contains all 13 serotypes included in PCV13 plus serotypes 22F and 33F. In the United States, serotypes 22F and 33F accounted for 15% of IPD in adults aged ≥65 years and 13% of IPD in adults aged 19 through 64 years with certain underlying medical conditions during 2018–2019 [9]. PCV20 contains all serotypes included in PCV13 and PCV15 (22F and 33F) as well as serotypes 8, 10A, 11A, 12F, and 15B. In the United States, these additional serotypes accounted for 27% of IPD in adults aged ≥65 years and 28% of IPD in adults with certain underlying medical conditions in 2018–2019 [9]. Phase II/III RCTs for PCV15 [10-12] and PCV20 [13, 14] showed immunogenicity and safety comparable with PCV13 or PPSV23; similar findings were observed for PCV15 use among adults with certain underlying medical conditions [15, 16].

A review of PCV13 and PPSV23 effectiveness against clinical outcomes was necessary for a few reasons: first, we expect that PCV15 and PCV20 will have similar effectiveness as PCV13 and PPSV23 and given these new vaccines were licensed based on safety and immunogenicity data only, this review would provide a comprehensive summary of overall pneumococcal vaccine effectiveness against clinical outcomes in adults. Second, PPSV23 was likely to be part of the new recommendations and previous literature found inconsistent evidence regarding PPSV23 effectiveness against non-bacteremic pneumococcal pneumonia. Lastly, PCV13 would be used as a comparator vaccine to the newer higher valent conjugate vaccines under discussion and a full review of PCV13 effectiveness against clinical outcomes was necessary. Therefore, we conducted a systematic review of literature on the efficacy or effectiveness of PCV13 and PPSV23 against VT-IPD and VT-pneumococcal pneumonia in adults to help inform the U.S. Advisory Committee on Immunization Practices’ (ACIP) discussions on the use of PCV15 and PCV20 among adults.

## Methods

### Literature search

In 2019, the Norwegian Public Health Institute (NIPH) conducted a systematic review of literature on pneumococcal vaccine effectiveness in adults for the World Health Organization’s Strategic Advisory Group of Experts [17]. We utilized the same search strategy (Appendix 1) and updated the search timeline to update the summary of evidence on pneumococcal vaccine efficacy or effectiveness on pneumococcal disease and mortality in adults.

An updated systematic literature search reviewed seven databases [Pubmed, Medline, Embase, CINAHL, Web of Science, Scopus (Web of Science), Epistemonikos, and Cochrane library] for relevant data published during April 2019 to February 2021. All relevant studies from the previous search by NIPH (published during January 2016 to March 2019) were also included in our review. Authors were contacted for additional information whenever possible. The results of all searches were entered into the Covidence systematic review software program [18].

The protocol for this review was developed in line with PRISMA-P recommendations [19] and registered with PROSPERO (CRD42021258668). The protocol was amended in February 2022 to include an additional year of search that was performed after the initial review. This search was conducted on April 4, 2022, to identify additional titles published during February 13, 2021, to March 15, 2022, to update the review.

### Inclusion and exclusion criteria

Data on PCV13 and PPSV23 efficacy or effectiveness against VT-IPD (defined as detection of *S. pneumoniae* in normally sterile sites, such as bacteremia and meningitis that are caused by serotypes contained in the vaccines) and VT-pneumococcal pneumonia (defined as pneumonia due to a *S. pneumoniae* serotype contained in the vaccines) were included.

We included published RCTs and observational studies (prospective and retrospective comparative cohort studies, case-control or nested case-control studies, indirect cohort studies, screening method studies) from middle- and high-income settings that evaluated direct effects of pneumococcal vaccination on adults (≥16 years). Systematic reviews on PCV13 or PPSV23 efficacy or effectiveness were included to leverage their bibliographies for relevant data. Only English language studies were included.

We excluded studies assessing vaccine impact, case reports, case-series, animal studies, modelling studies, health economic evaluations, pneumococcal carriage studies, narrative reviews, studies that only included data on vaccines that are not currently used in the United States (e.g., PCV7, PCV10),, studies evaluating indirect effects of pediatric pneumococcal vaccination on adult populations, and studies specifically targeting adults with immunocompromising conditions. In addition, because pneumococcal disease epidemiology differs from that in the United States, we excluded studies conducted in low-income settings.

### Data abstraction and cleaning

Titles and abstracts of all citations identified through the literature search were independently screened for relevance and inclusion by two reviewers using Covidence [20]. Articles meeting inclusion through title and abstract screening were then double screened through full-text review. Discordant reviews during title and abstract or full-text review were blindly reviewed for inclusion by an independent third reviewer.

Data were independently abstracted by two reviewers for study information (study characteristics, methods, results) using standardized data collection tools and entered into a Microsoft Excel spreadsheet.

Data were also assessed for quality of evidence. Randomized studies were assessed for quality using the Cochrane Risk of Bias 2.0 tool [21], which included consideration of the appropriate generation of random allocation sequences; concealment of the allocation sequences; blinding of participants, healthcare providers, data collectors, and outcome adjudicators; and proportion of patients lost to follow-up. Non-randomized studies were assessed considering the data elements included in the Newcastle-Ottawa scale (NOS) [22] to address potential sources of bias in cohort and case-control studies. Each study was assessed for risk of bias independently by two reviewers. A third reviewer was consulted when there was discordance of overall quality of evidence between the two independent reviewers. Studies with scores ≥7 (out of 9) were deemed high quality/low risk of bias, scores between 4 to 6 (out of 9) were deemed medium quality/some risk of bias, and scores ≤3 were deemed low quality/high risk of bias.

### Data analysis

We conducted descriptive analyses of studies stratified by outcome (VT-IPD or VT-pneumococcal pneumonia), vaccine product (PCV13 or PPSV23), and study design (RCT or observational study). When feasible, we conducted meta-analyses by outcome, product, and study design using Comprehensive Meta-Analysis Version 3 [23]. Random effects models were used to pool estimates by stratum. Because PPSV23 has been used for adults much longer than PCV13, time since PPSV23 vaccination was more variable across studies compared with PCV13 studies. To account for waning of vaccine effectiveness over time [24-29], we limited the PPSV23 analysis to data from individuals who were vaccinated within 5 years of assessment. When data were available, we performed sub-analyses by underlying conditions and limiting to a younger age group of adults aged 65 through 74 years to account for heterogeneity in the study population between observational studies.

## Results

### Literature Search

The literature search yielded a total of 5,085 articles, 2,499 from the NIPH search that covered titles published during January 2016 to April 2019 and 2,586 from the updated search that covers titles published during April 2019 to March 2022. From the updated search, 1,764 studies were screened for eligibility; 1,525 were excluded during title and abstract screening. During the full-text review, 239 articles were screened for eligibility; 229 were excluded. Ten studies were included from the updated searches and nine studies were included from the NIPH search, totaling 19 studies for inclusion in the analysis (Figure 1). Seven studies evaluated PCV13 and 13 evaluated PPSV23; one study evaluated both PCV13 and PPSV23 against VT-pneumococcal pneumonia [30]. Efficacy or effectiveness against VT-IPD was evaluated in 13 studies and against VT-pneumococcal pneumonia in eight studies; two studies evaluated pneumococcal vaccine effectiveness against both VT-IPD and VT-pneumococcal pneumonia (Table) [5, 31].

**Table.**
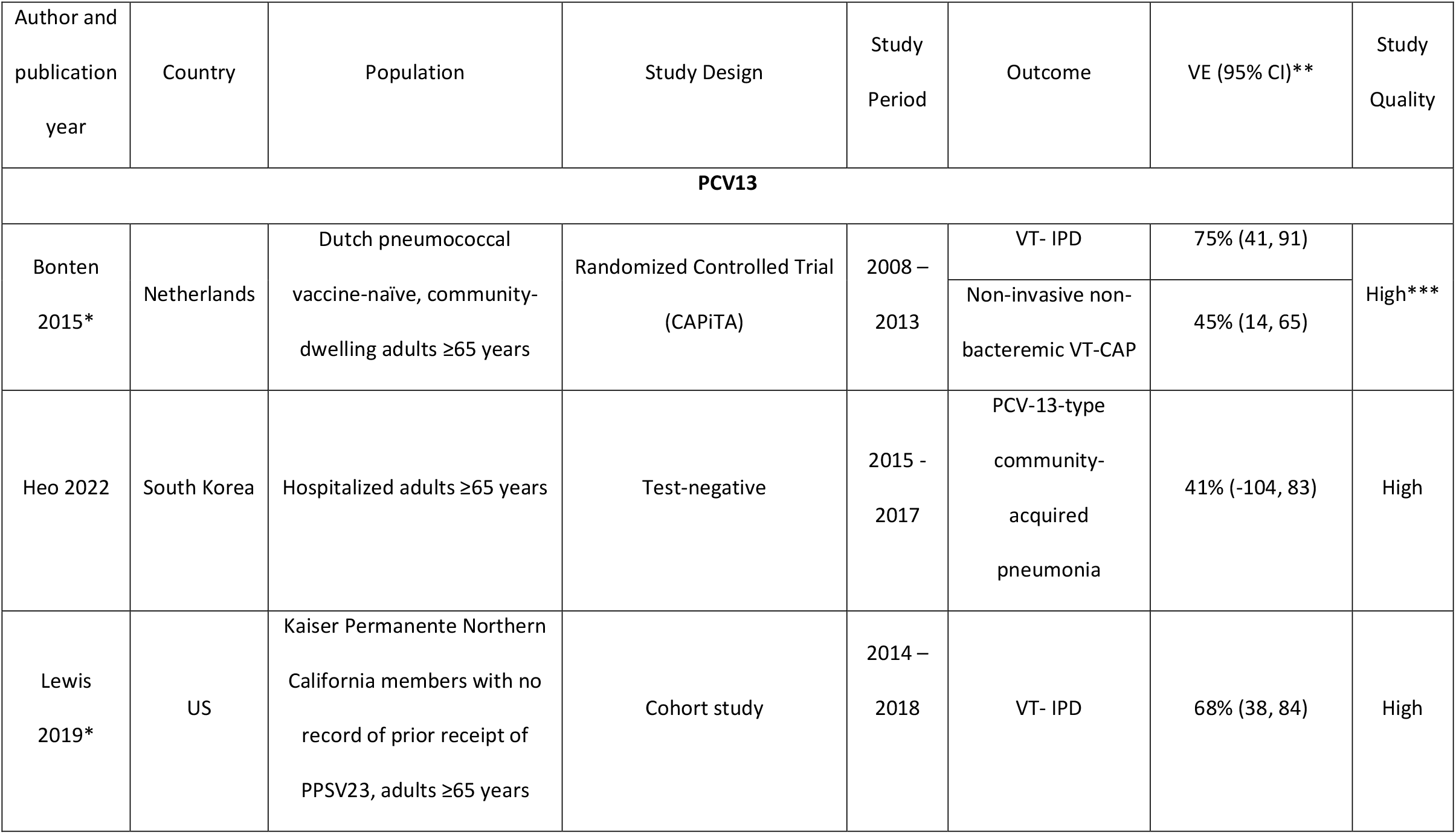
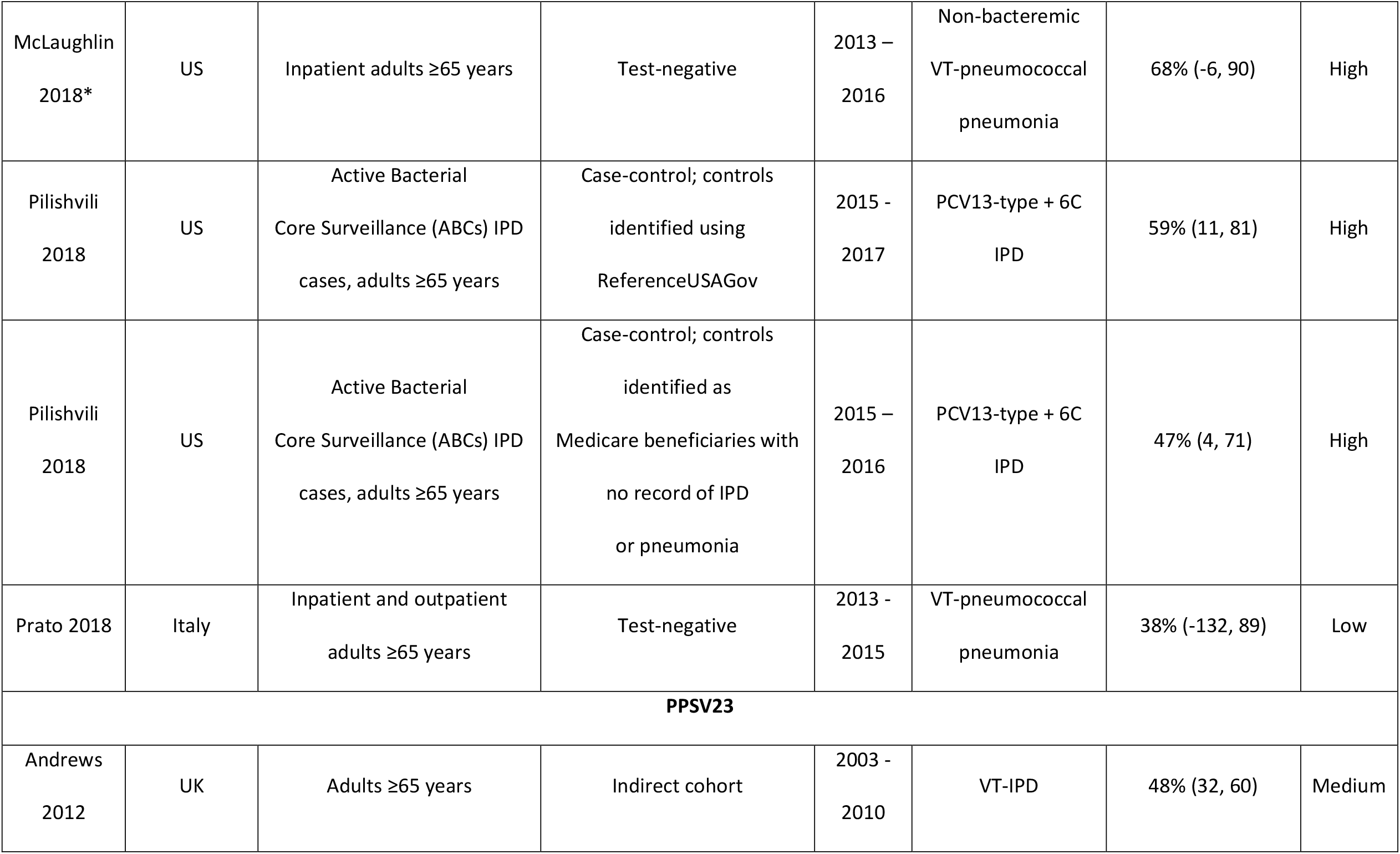
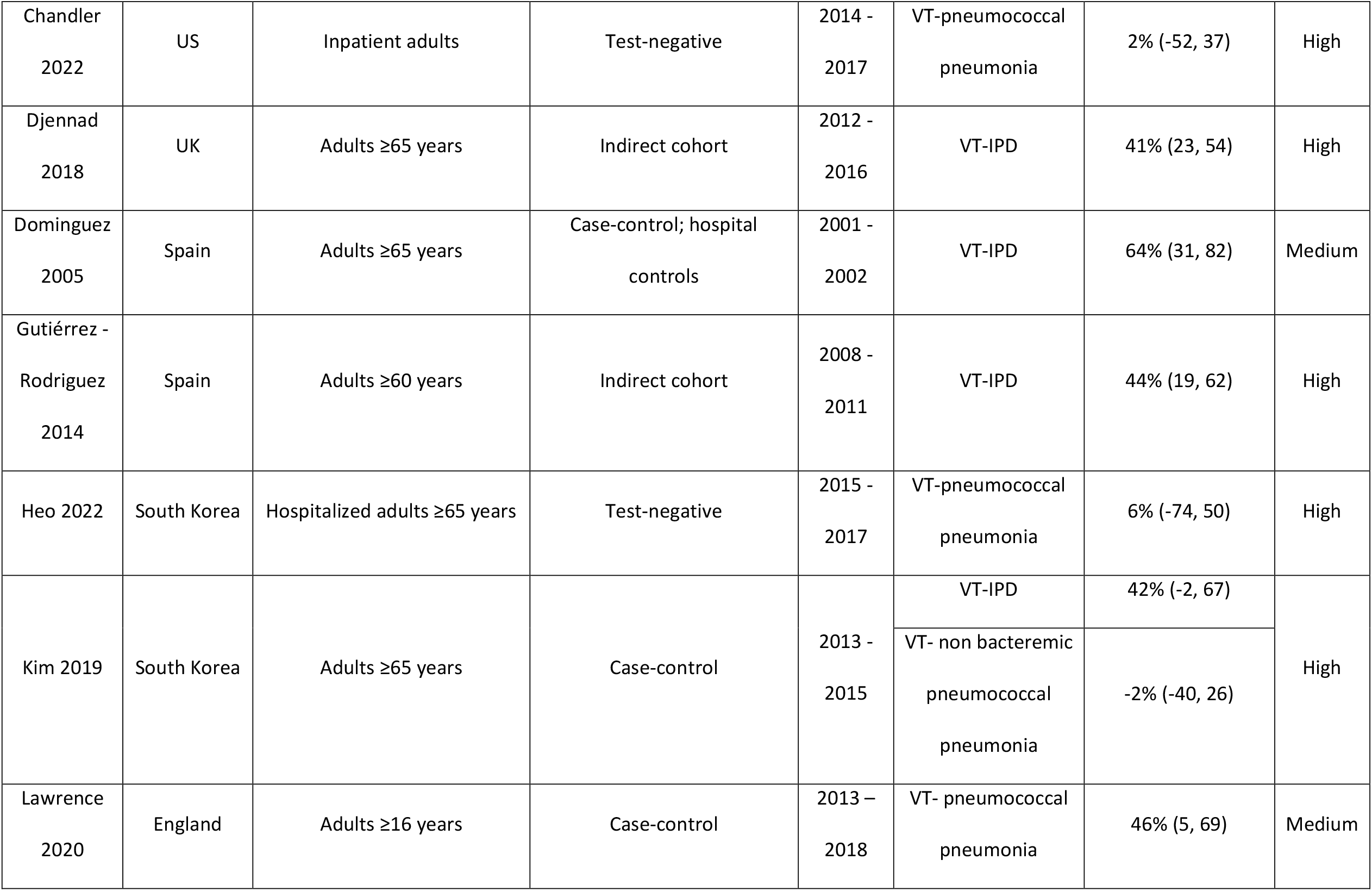
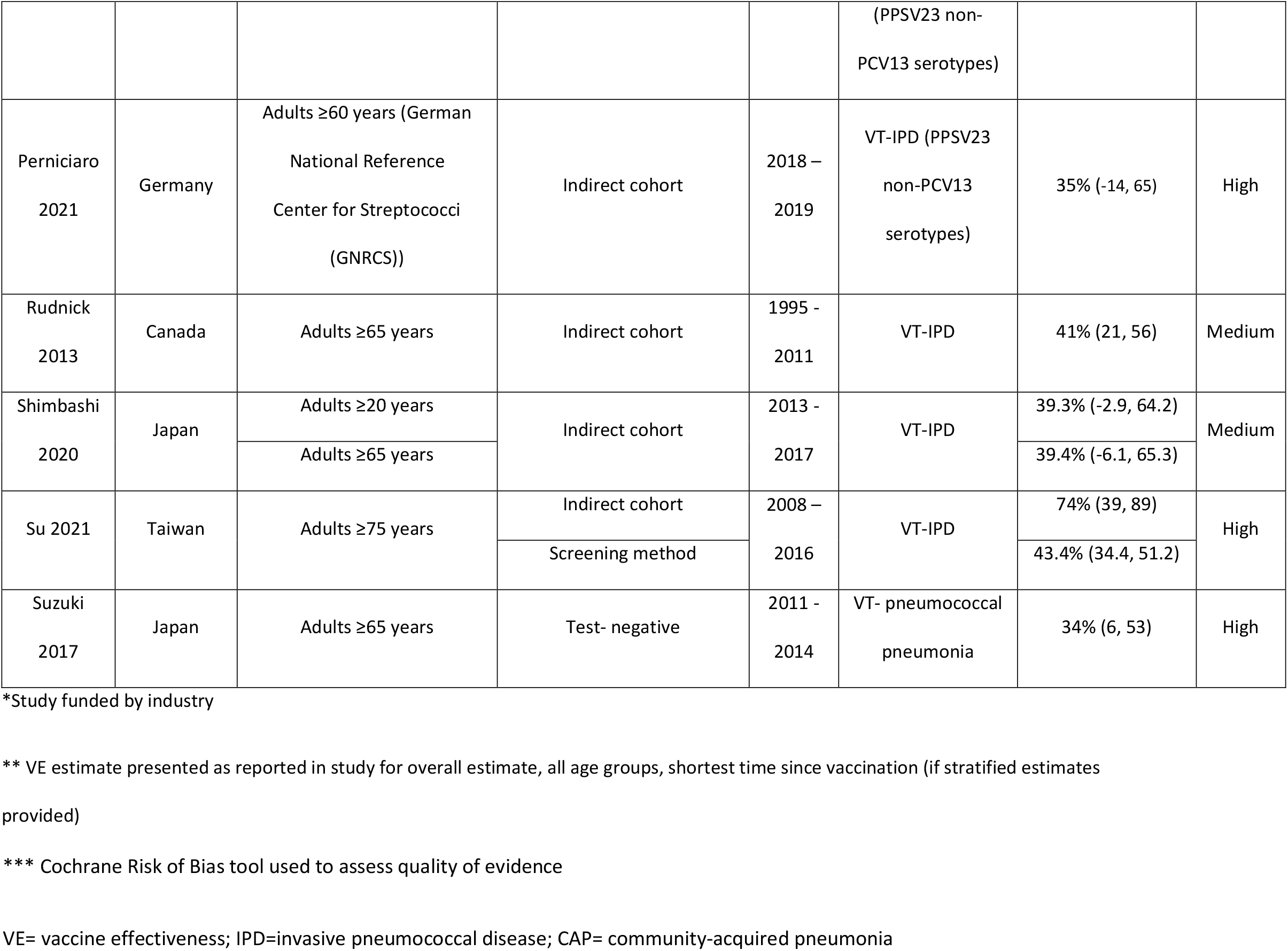
Characteristics of Included Pneumococcal Vaccine Effectiveness Studies.

**Figure 1.**
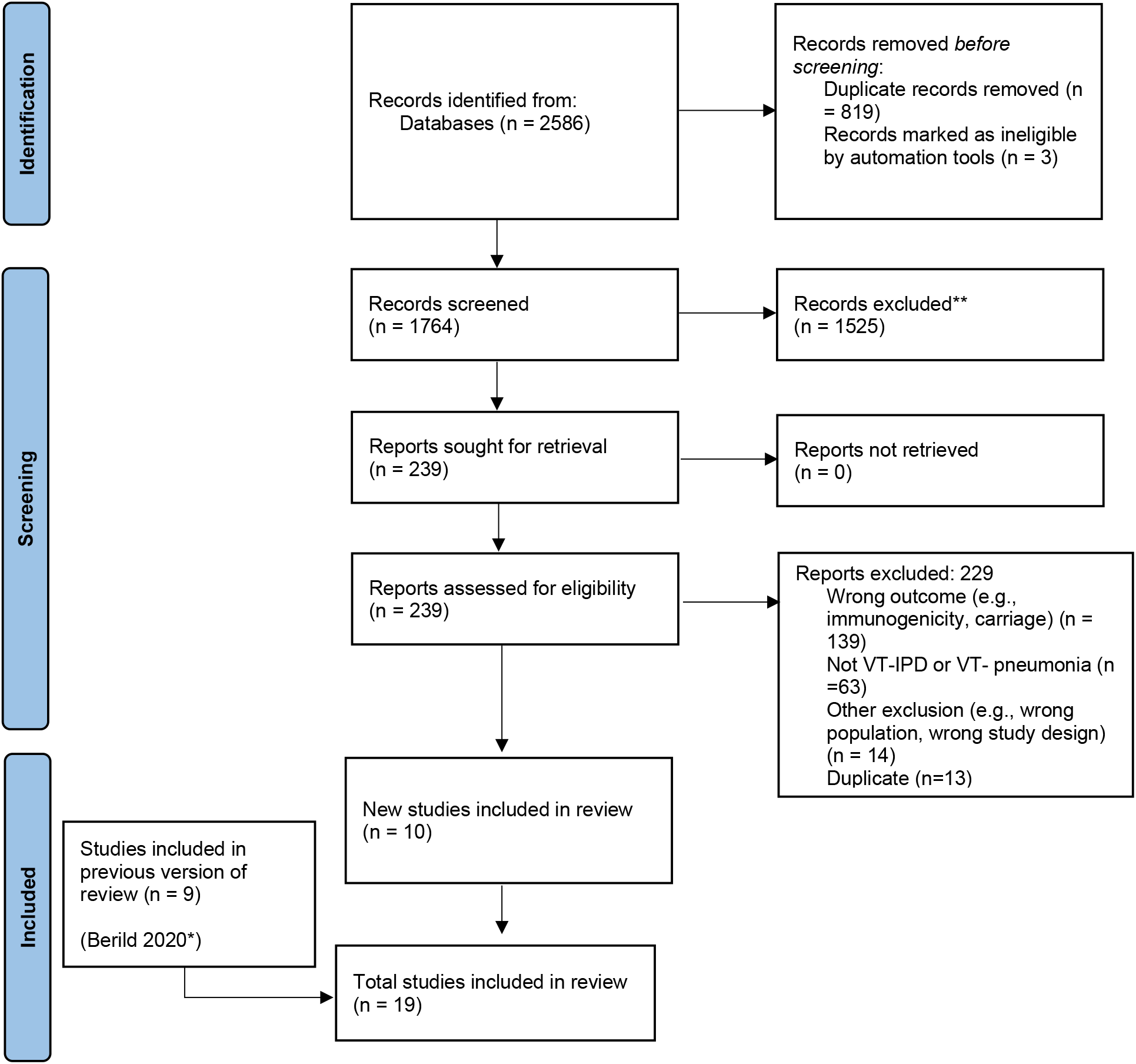
PRISMA Study flow diagram. *Berild 2020 is the publication for the Norwegian Public Health Institute (NIPH) systematic review of literature on pneumococcal vaccine effectiveness in adults conducted for the World Health Organization’s Strategic Advisory Group of Experts.

### Vaccine-type invasive pneumococcal disease

#### PCV13

We identified four studies evaluating PCV13 efficacy or effectiveness against VT-IPD. Only one RCT was identified, with a reported efficacy of 75% (95% CI: 41%, 91%) against VT-IPD in Dutch pneumococcal vaccine-naïve, community-dwelling adults aged ≥65 years [5]. Vaccine effectiveness (VE) estimates from three observational studies ranged from 47%–68% against VT-IPD in adults aged ≥65 years [32-34] (Table 1). Lewis et al evaluated PCV13 VE in a cohort of U.S. adults aged ≥65 years who were enrolled in Kaiser Permanente Northern California insurance and had not received PPSV23 before age 65 years. PCV13 VE was 68% (95% CI: 38%, 84%) against all VT-IPD, and 53% (95% CI: -10%, 80%) against serotype 3 IPD; the study reported an increase in the incidence of IPD during the study period (2014–2018), largely due to serotype 3. The other two observational studies by Pilishvili et al were case-control studies that used Active Bacterial Core surveillance data to identify cases, but different sources to identify controls [33, 34]. The VE estimate for PCV13 against IPD caused by PCV13 + 6C serotypes was 59% (95% CI: 11%, 81%) using controls identified from the commercial database ReferenceUSAGov (InfoGroup) [33] and 47% (95% CI: 4%, 71%) using Medicare beneficiaries as controls; this estimate increased to 67% (95% CI: 11%, 88%) when excluding serotype 3 IPD cases [34]. Due to overlap in cases between the two Pilishvili et al studies, a pooled VE was not estimated between studies.

#### PPSV23

We did not identify any RCTs assessing PPSV23 efficacy against VT-IPD. Nine observational studies reported VE among those who received PPSV23 within 5 years of the study assessment [24-26, 28, 31, 35-38]. Of these, seven were indirect cohorts and two were case-control studies. Studies evaluated VE among adults aged ≥ 60 years from Canada, Germany, United Kingdom (n=2), South Korea, Japan, Taiwan, and Spain (n=2). Of note, Perniciaro et al reported a -30% (95% CI: -159%, 31%) PPSV23 effectiveness against VT-IPD; however, the authors observed that vaccination rates were low (16%) among adults who had received PPSV23 within five years prior to the study. Additionally, over 20% of cases were caused by serotype 3. Therefore, we report the VE against PPSV23/non-PCV13 serotype IPD, which was 35% (95% CI: -14%, 65%) [38]. The pooled VE estimate from these nine observational studies was 45% (95% CI: 37%, 51%; I^2^=0%) (Figure 2).When limited to three studies in adults without immunocompromising conditions [24, 25, 28], the pooled VE estimate was 60% (95% CI: 47%, 69%; I^2^= 0%) (Figure 3). The pooled VE estimate for a sub-analysis restricting to adults aged 65 through 74 years [24, 25, 31] was 52% (95% CI: 36%, 64%; I^2^= 5.2%) (Figure 4).

**Figure 2.**
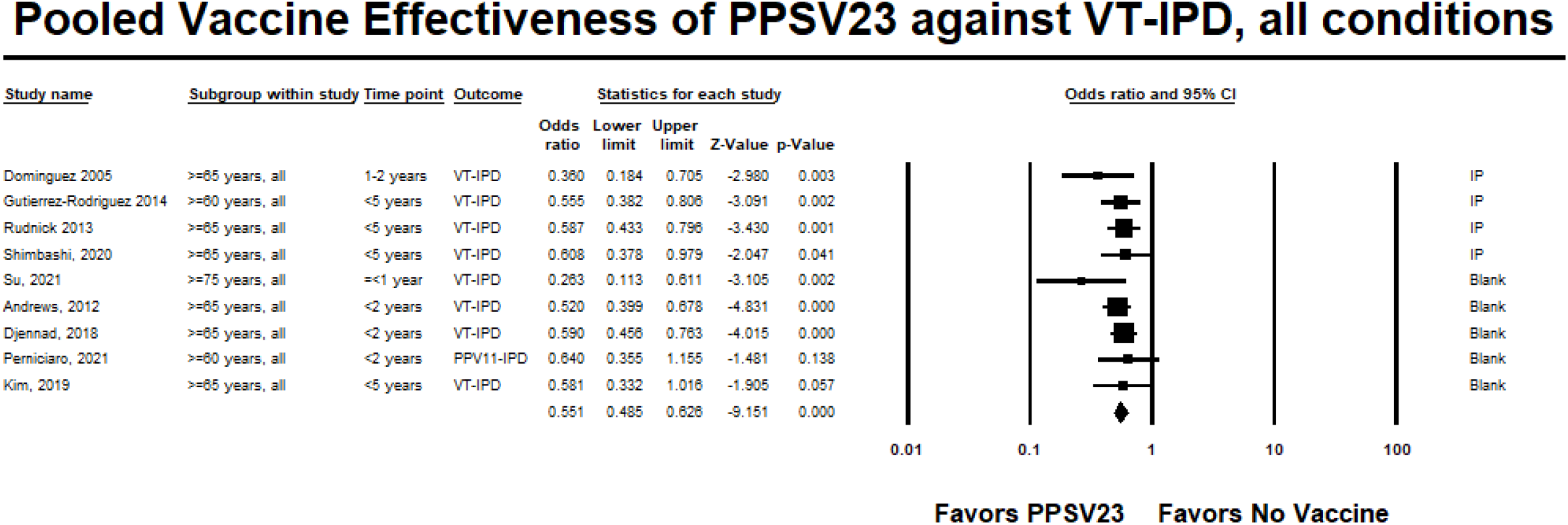
**Pooled Vaccine Effectiveness Estimate of 23-Valent Pneumococcal Polysaccharide Vaccine Against Vaccine-Type Invasive Pneumococcal Disease in Adults Aged ≥ 60 Years: Observational studies, <5 years since vaccination**

**Figure 3.**
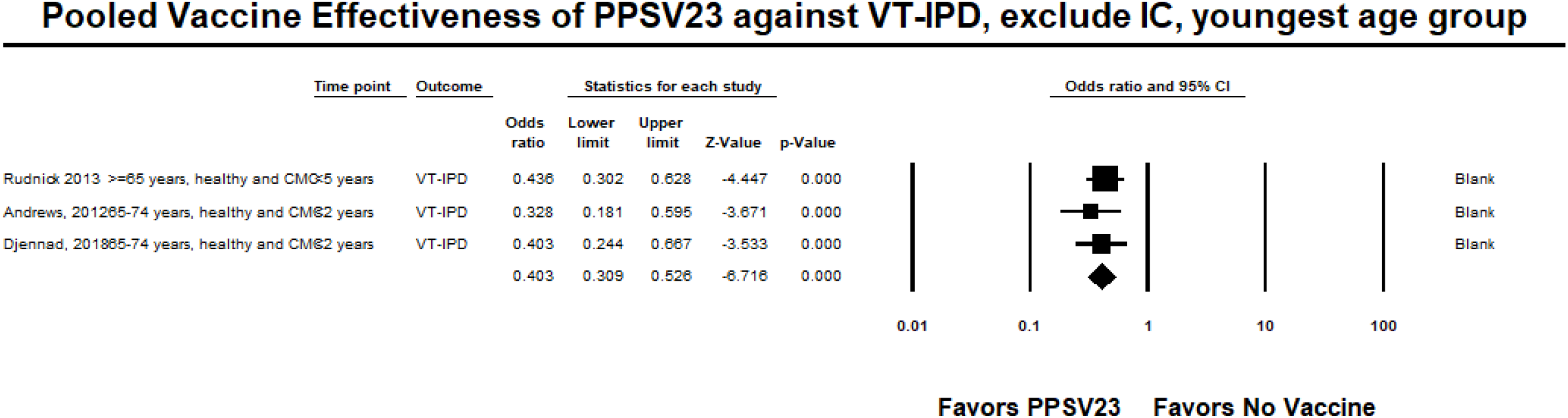
**Pooled Vaccine Effectiveness of 23-Valent Pneumococcal Polysaccharide Vaccine Against Vaccine-Type Invasive Pneumococcal Disease in Healthy Adults: Observational studies, <5 years since vaccination**

**Figure 4.**
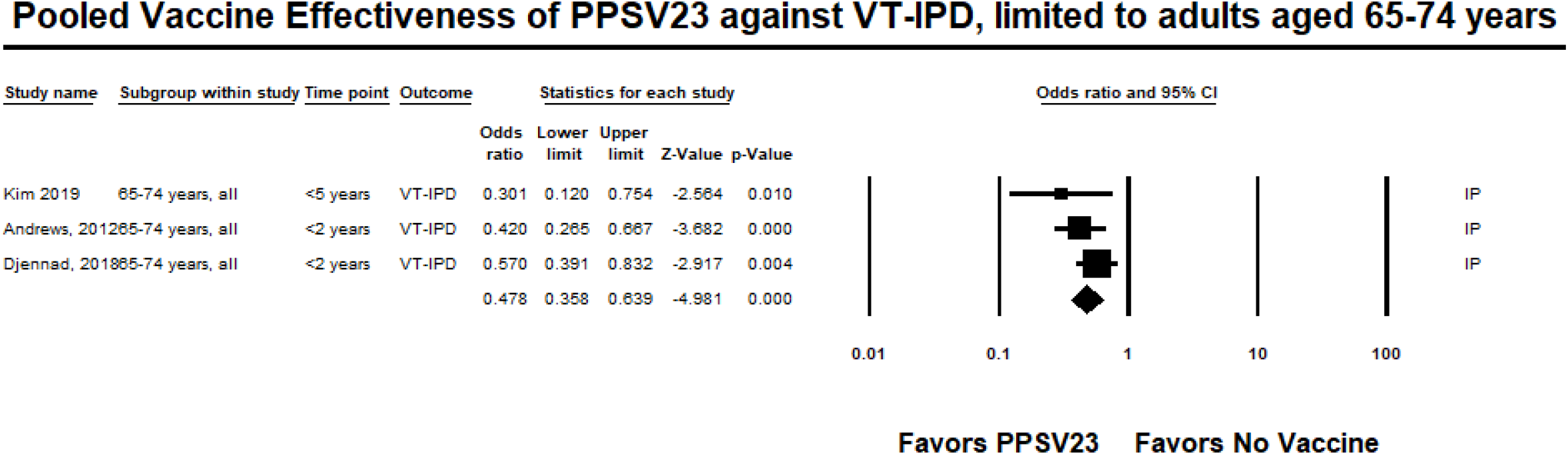
**Pooled Vaccine Effectiveness of 23-Valent Pneumococcal Polysaccharide Vaccine Against Vaccine-Type Invasive Pneumococcal Disease in Adults Aged 65–74 years: Observational studies, <5 years since vaccination**

### Vaccine-type Pneumococcal Pneumonia

#### PCV13

We identified four studies evaluating PCV13 efficacy or effectiveness against VT-pneumococcal pneumonia. One RCT, among Dutch adults aged ≥65 years, reported an efficacy of 45% (95% CI: 14%, 65%) against the first episode of VT-nonbacteremic and noninvasive pneumococcal pneumonia [5]. In addition to the RCT, we identified three test-negative design (TND) observational studies for PCV13. The study by Prato et al was conducted among outpatient and inpatient Italian adults aged ≥65 years living in the Apulia region [39]. Many participants had one or more underlying comorbidity including chronic heart disease (53%), chronic respiratory disease (44%), and diabetes (25%). Patients were considered to have pneumococcal pneumonia if they had a positive PCR result for *S. pneumoniae* from blood, sputum, or bronchoalveolar-lavage. Prato et al reported a crude VE of 38% (95% CI: -132%, 89%) against VT-pneumococcal pneumonia, and an adjusted VE was not reported [39]. The study by McLaughlin et al was conducted as a nested study within a larger population-based surveillance study of U.S. adults in Kentucky. Adults aged ≥65 years hospitalized with community acquired pneumonia (CAP) were considered for enrollment. Most participants (88%) had one or more at-risk (defined as immunocompetent, but presence of a chronic disease such as congestive heart failure, diabetes mellitus, or liver disease) or high-risk (defined as immunocompromised) condition, and almost half of participants were considered high-risk (46%). The adjusted VE was 68% (95% CI: -6%, 90%) against nonbacteremic VT-pneumococcal pneumonia [40]. Heo et al evaluated the effectiveness of PCV13, PPSV23, and sequential PCV13/PPSV23 vaccination against pneumococcal pneumonia among older South Korean adults aged ≥65 years hospitalized with CAP. The mean age of participants was 76.7 ± 6.9 years and many participants had ≥1 underlying comorbidity (80%) or ≥2 underlying comorbidities (45%). The adjusted VE against VT-pneumococcal pneumonia was 41% (95% CI: -104%, 83%) for all adults aged ≥65 years, [30]. Pooled VE of the TND observational studies was not estimated since Prato et al only reported a crude VE estimate.

#### PPSV23

We did not identify any RCTs evaluating efficacy of PPSV23 against VT-pneumococcal pneumonia. We identified five observational studies with VE estimates ranging from –2% to 46% [29-31, 41, 42]. The study by Kim et al was a hospital-based case-control study conducted among South Korean adults aged ≥65 years. The median age of the study population was 76 years and many participants had one or more underlying medical conditions such as chronic pulmonary disease (37% of cases, 23% of controls) or diabetes mellitus (24% of cases, 30% of controls). Approximately 26% of cases and 37% of controls had an immunocompromising condition. The adjusted VE against VT-nonbacteremic pneumococcal pneumonia was -2% (95% CI: -40%, 26%) [31]. The four remaining observational studies were TND. Lawrence et al used a nested TND of a prospective cohort study of adults aged ≥16 years hospitalized with CAP at two hospitals in England. The mean age was similar between cases and controls (66.5 versus 65.4 years). The most common comorbidities among cases and controls were chronic obstructive pulmonary disease (COPD) (24% for cases and controls), chronic lung disease (27% for cases and controls), and hypertension (24% of cases, 25% of controls). The adjusted VE of PPSV23 against VT-pneumococcal pneumonia (non-bacteremic and bacteremic) among those vaccinated <5 years was –7% (95% CI: -54%, 26%). Lawrence et al observed that serotype 5 caused the largest proportion of cases of PPSV23 serotype pneumonia <5 years since vaccination and considered the potential cross-reactivity with other streptococcal strains that express serotype 5; however, the authors noted the potential cross-reactivity did not alone explain the difference in effects among vaccinated and unvaccinated patients. Therefore, we report the VE against PPSV23/non-PCV13 serotypes pneumococcal pneumonia, which was 46% (95% CI: 5%, 69%) [41]. Suzuki et al conducted a prospective TND study among adults aged ≥65 years at four community-based hospitals in Japan. Presence of underlying medical conditions was similar between cases and controls. The adjusted VE of PPSV23 against VT-pneumococcal pneumonia among outpatients and inpatients was 34% (95% CI: 6%, 53%) [29]. Heo et al, described above, reported an adjusted VE of 6% (95% CI: -74%, 50%) against VT-pneumococcal pneumonia among participants aged ≥65 years [30]. Chandler et al conducted a TND study among hospitalized CAP patients identified through the University of Louisville Pneumonia Study database that was also used for the study by McLaughlin et al [40]. The median age of vaccinated patients was 68 years, compared to 65 years among non-vaccinated patients. Vaccinated patients were more likely to have comorbid conditions such as COPD (58% among vaccinated patients versus 47% among unvaccinated patients), renal failure (33% versus 26%), or diabetes (37% versus 31%). Chandler et al reported 2% (95% CI: -52%, 37%) vaccine effectiveness against VT-pneumococcal pneumonia in adults aged ≥65 years who received PPSV23 within 5 years of assessment. [42]. Pooled VE of PPSV23 against VT-pneumococcal pneumonia from the five observational studies was 18% (95% CI: -4%, 35%; I^2^=0%) (Figure 5). When limited to adults aged 65 through 74 years, the pooled VE estimate from three studies [29-31] was 26% (95% CI: -6%, 49%; I^2^=0%) (Figure 6).

**Figure 5.**
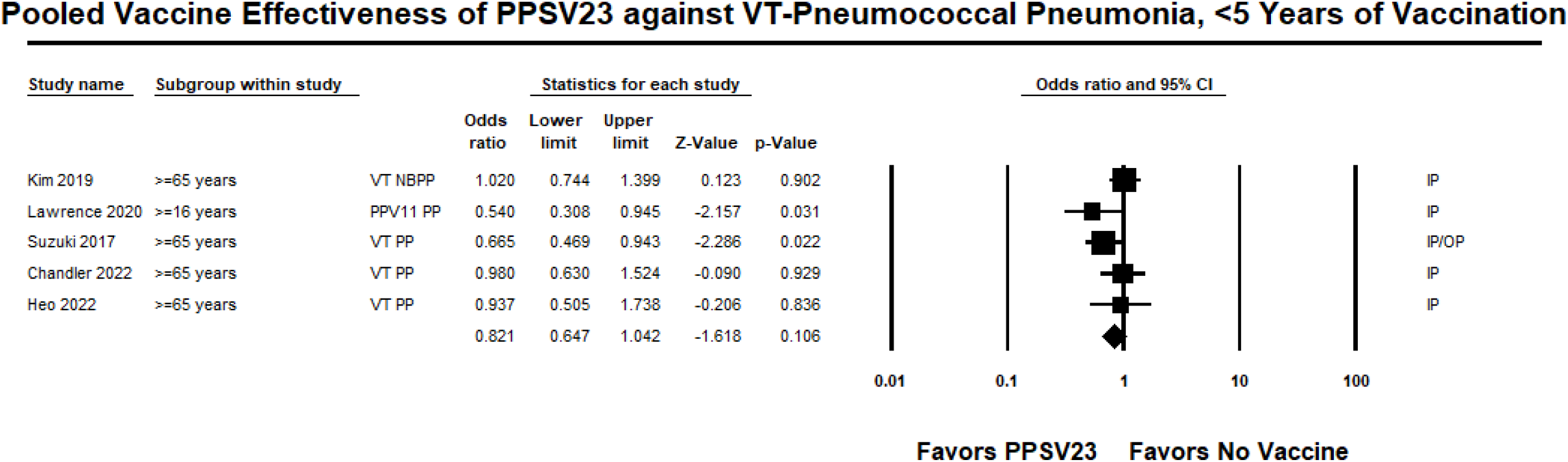
**Pooled Vaccine Effectiveness of 23-Valent Pneumococcal Polysaccharide Vaccine Against Vaccine-Type Pneumococcal Pneumonia in Adults aged ≥16 Years: Observational studies, <5 years since vaccination**

**Figure 6.**
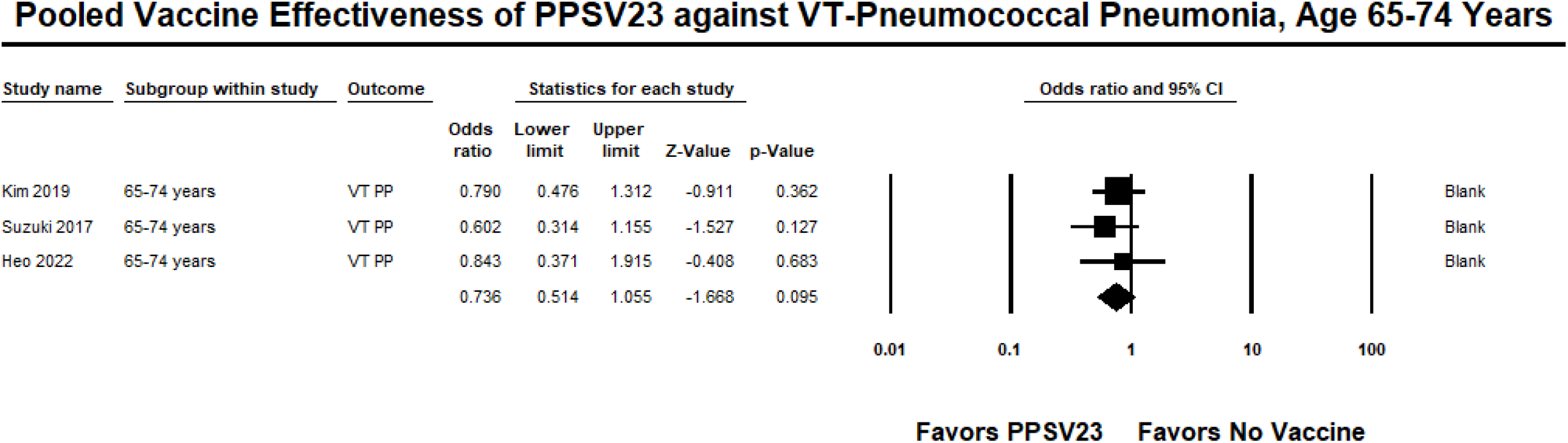
**Pooled Vaccine Effectiveness of 23-Valent Pneumococcal Polysaccharide Vaccine Against Vaccine-Type Pneumococcal Pneumonia in Adults Aged 65 – 74 Years: Observational studies, <5 years since vaccination**

### Quality of Evidence

The risk of bias for the one RCT [5] was low; therefore, the quality of evidence was high for this study. Regarding observational studies, scores using the Newcastle-Ottawa scale ranged from 3 to 8 (out of 9) for PCV13 studies; three studies were deemed high quality (score of 7 or 8). Prato et al scored 3 for lack of information regarding selection of controls and exposure criteria as well as lack of controlling for potential confounders; it was rated low for quality of evidence. For PPSV23 observational studies, eight studies were deemed high quality (score of 7 to 8) [25, 26, 29-31, 37, 38, 42] and five were deemed medium quality (score of 4 to 6) [24, 28, 35, 36, 41]. All studies that scored medium quality lacked information regarding source of controls or ascertainment of exposure (i.e., how vaccination history was obtained).

## Discussion

Recent data continue to support that both PCV13 and PPSV23 are effective against VT-IPD in adults. However, there are a few differences in the published studies that warrant additional discussion. When considering PCV13, the VE estimates from Pilishvili et al were lower than the estimates from the RCT; this is expected, as effectiveness estimates from observational studies are often lower than efficacy estimates from clinical trials, which occur in a more controlled setting. Furthermore, the lower observed estimates could be due to differences in the study population. Specifically, 54–60% of cases and 31–32% of controls in the case-control studies by Pilishvili et al had immunocompromising conditions, whereas adults with immunocompromising conditions at enrollment were not included in the RCT. Regarding PCV13 effectiveness against VT-pneumococcal pneumonia, findings from observational studies supported findings from the RCT, which showed 45% effectiveness. However, estimates from the observational studies had wide confidence intervals.

When considering PPSV23, the pooled VE estimate for PPSV23 against VT-IPD was slightly lower than what was observed for PCV13, even after limiting the data to <5 years since vaccination. These findings are not unexpected as immunogenicity studies have observed polysaccharide vaccines to be less immunogenic compared to conjugate vaccines. This review reinforced the existing evidence for PPSV23 effectiveness against VT-IPD.

Until recently, there were limited data specifically assessing PPSV23 VE against VT-pneumococcal pneumonia. Previous studies that reported PPSV23 effectiveness against pneumococcal pneumonia or CAP had variable results [43-45]. A review and meta-analysis by Falkenhorst et al estimated a pooled VE against pneumococcal pneumonia of 64% (95%CI: 35– 80%) in clinical trials and 48% (95%CI: 25–63%) in cohort studies, after excluding studies with high risk of bias [43]. Conversely, a review by Schiffner-Rohe et al reported that three of four studies showed no efficacy against pneumococcal pneumonia [44]. Recent studies assessing PPSV23 effectiveness against VT-pneumococcal pneumonia allowed us to conduct a pooled VE estimate, which suggests that PPSV23 provides some protection against VT-pneumonia within 5 years of vaccination.

In October 2021, based in part on this analysis, the U.S. ACIP voted to recommend PCV15 in series with PPSV23 or PCV20 use in U.S. adults who have not previously received a PCV [9]. This recommendation simplified previous recommendations on pneumococcal vaccine use in adults, which differed by age- and risk-groups [6]. The new recommendations also provide an opportunity for broader PCV coverage among adults compared to previous recommendations, in which routine PCV use was only recommended for adults with immunocompromising conditions. Because licensure of PCV15 and PCV20 was based on immunogenicity and safety data only, there are remaining questions regarding clinical implications of implementing these vaccines. For example, in clinical trials, PCV20 elicited slightly lower immune responses compared with PCV13 for the 13 shared serotypes, although noninferiority criteria were met [13]. The clinical implications of these findings, especially in adults with immunocompromising conditions who were not included in Phase II and III trials, are unknown. Furthermore, it is unknown whether PCV15 use in series with PPSV23 will provide better protection against the 15 shared serotypes compared to PCV20 only. A study from Korea reported that among adults aged 65 through 74 years, PCV13 in series with PPSV23 might result in improved protection against non-bacteremic VT-pneumococcal pneumonia compared with PCV13 or PPSV23 alone [30].

There are limitations to our review. We identified only one RCT that evaluated PCV13 [5] and none that evaluated PPSV23 efficacy against VT-pneumococcal disease in adults. RCTs provide the most rigorous evidence of vaccine efficacy. However, we identified several observational studies evaluating both PCV13 and PPSV23 effectiveness against VT-IPD and VT-pneumonia. Heterogeneity in study populations, e.g., different age distributions and different proportions of people with underlying conditions included, for PCV13 and PPSV23 studies may have affected the VE estimates. Furthermore, these differences in populations may in part contribute to the range of results observed across studies, though we have attempted to address this limitation by conducting stratified analysis of the data. Despite these limitations, this was an exhaustive and systematic review of literature on pneumococcal vaccine products for adult use and provides a comprehensive, current landscape of the evidence regarding VE for PCV13 and PPSV23 against VT-IPD and VT-pneumococcal pneumonia in adults.

In conclusion, our review showed that PCV13 and PPSV23 are both effective against VT-IPD and VT-pneumococcal pneumonia in adults. These data helped inform decisions on PCV15 and PCV20 use in adults in the United States, which currently do not have efficacy or effectiveness data against clinical outcomes and could be used to inform similar decision in other countries. Post-licensure effectiveness and impact studies of PCV15 and PCV20 on pneumococcal disease in adults are needed to assess the impact of the new recommendations.

## Data Availability

All relevant data are within the manuscript and its Supporting Information files.

## Appendix 1. Systematic literature search strategy

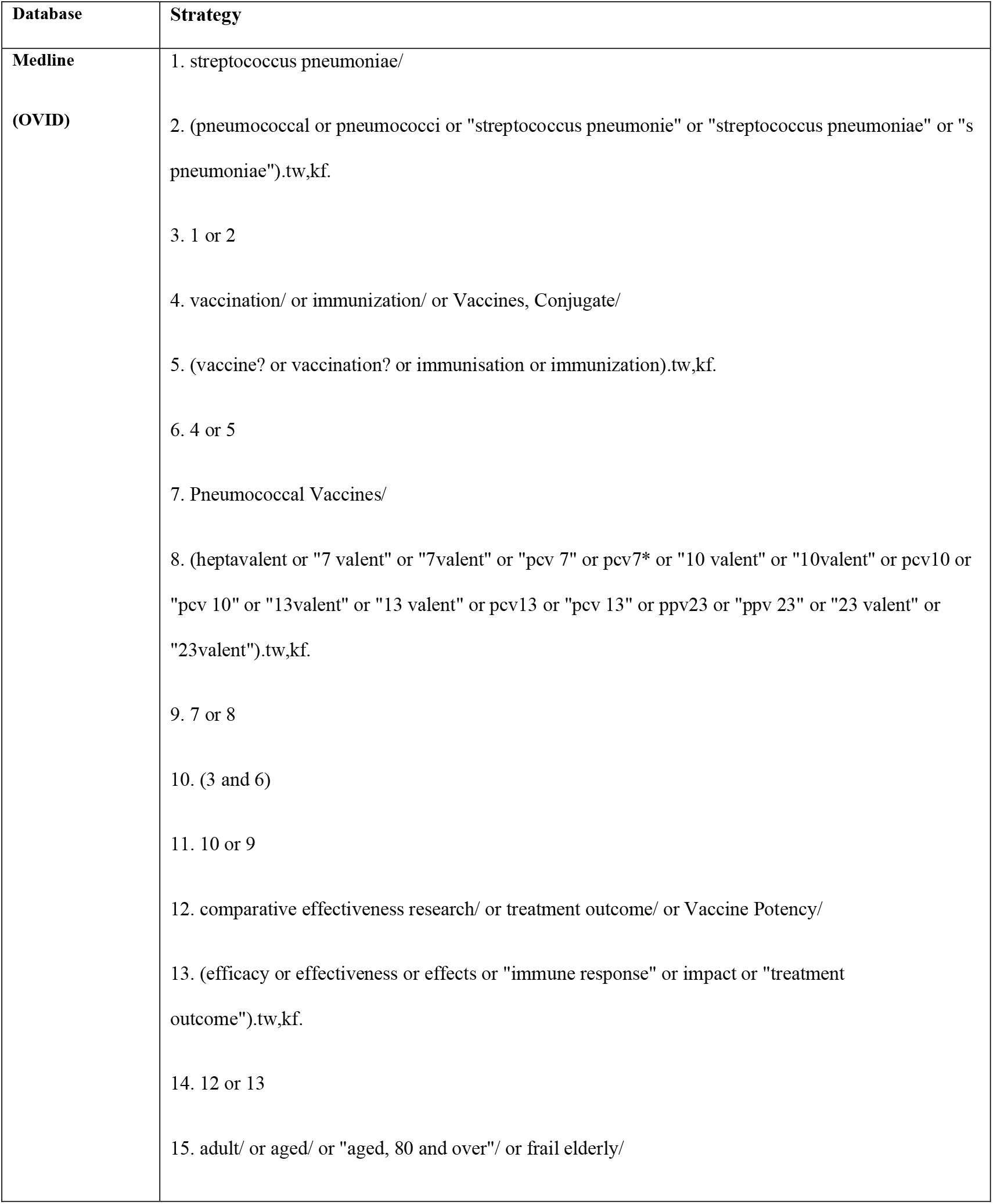
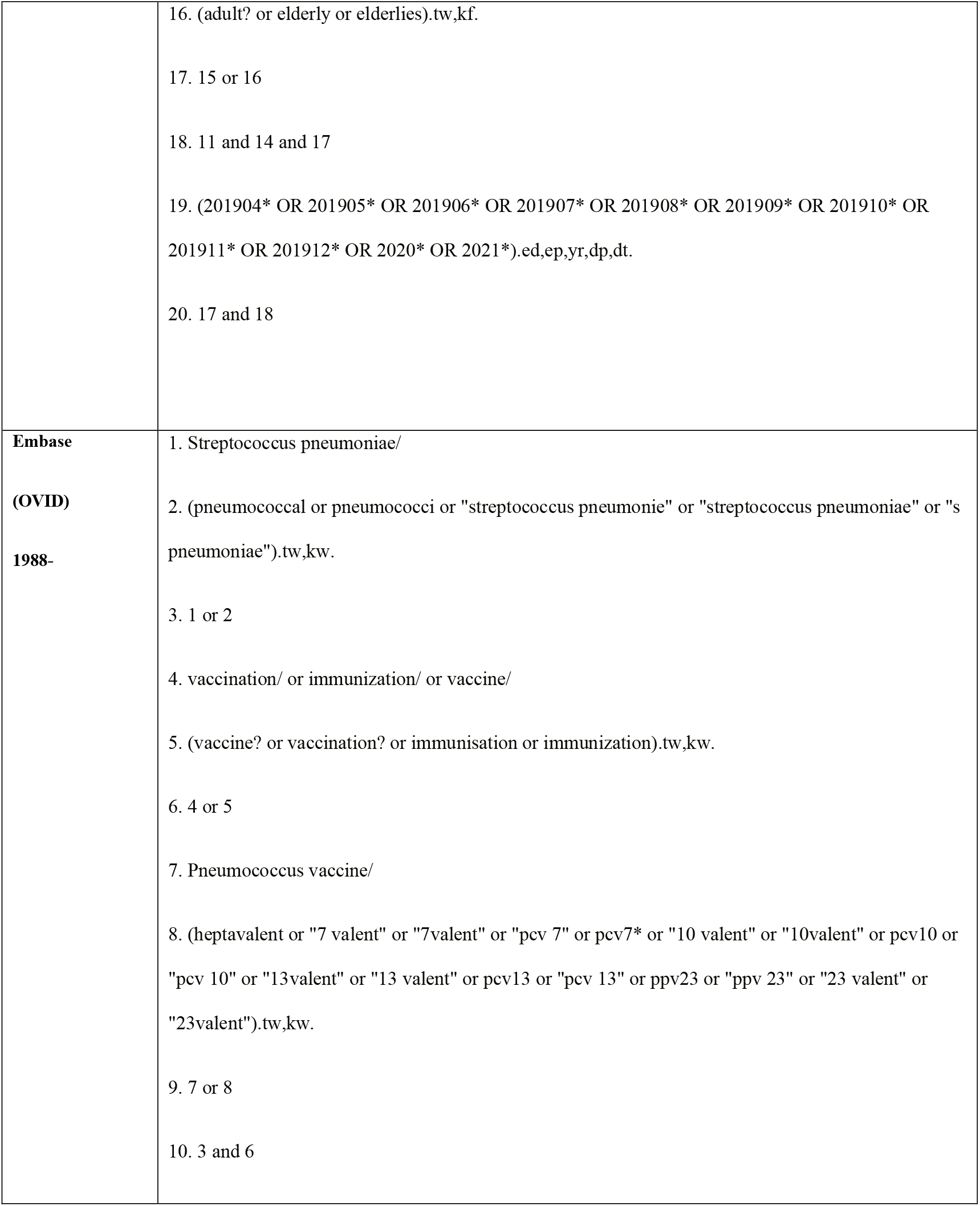
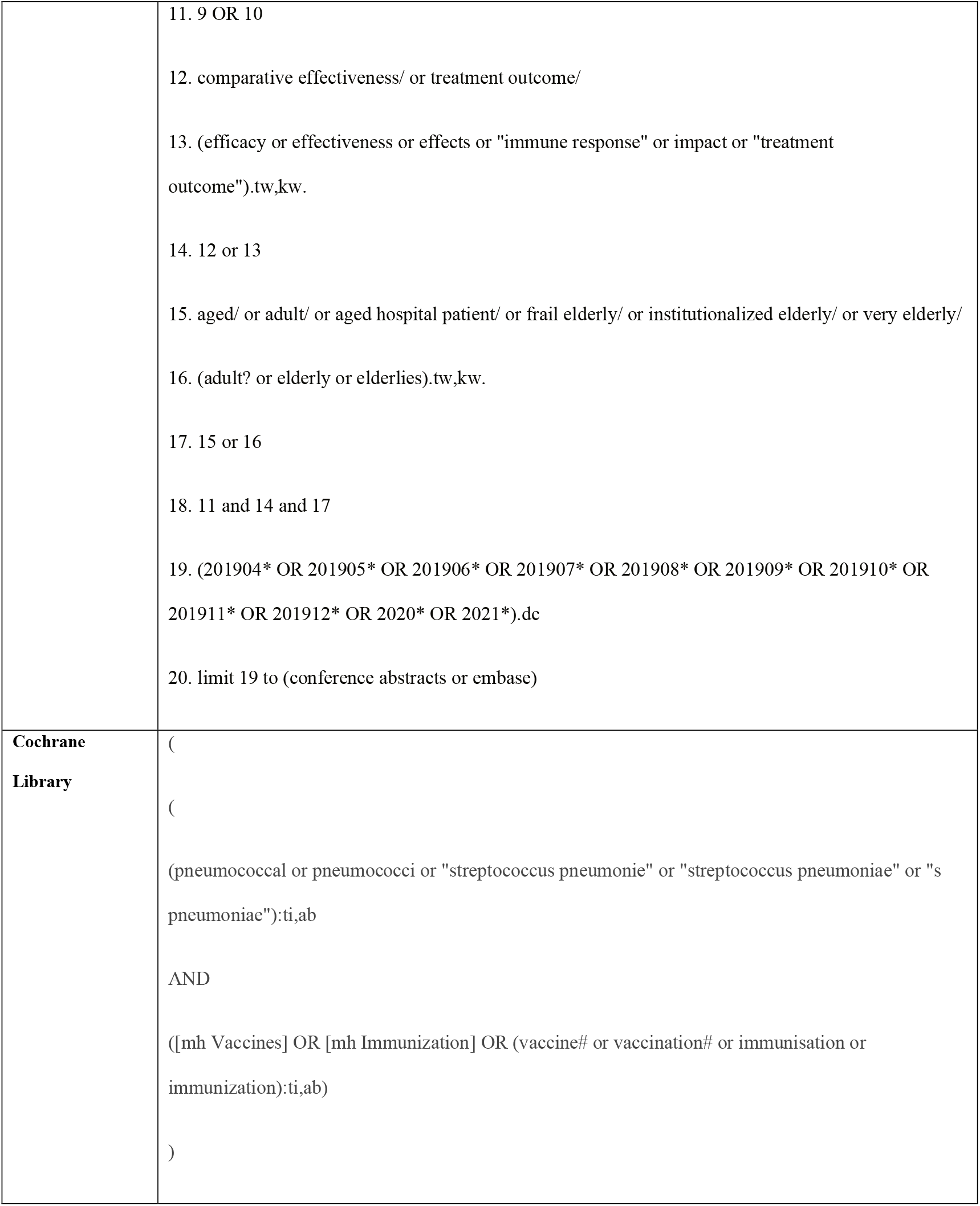
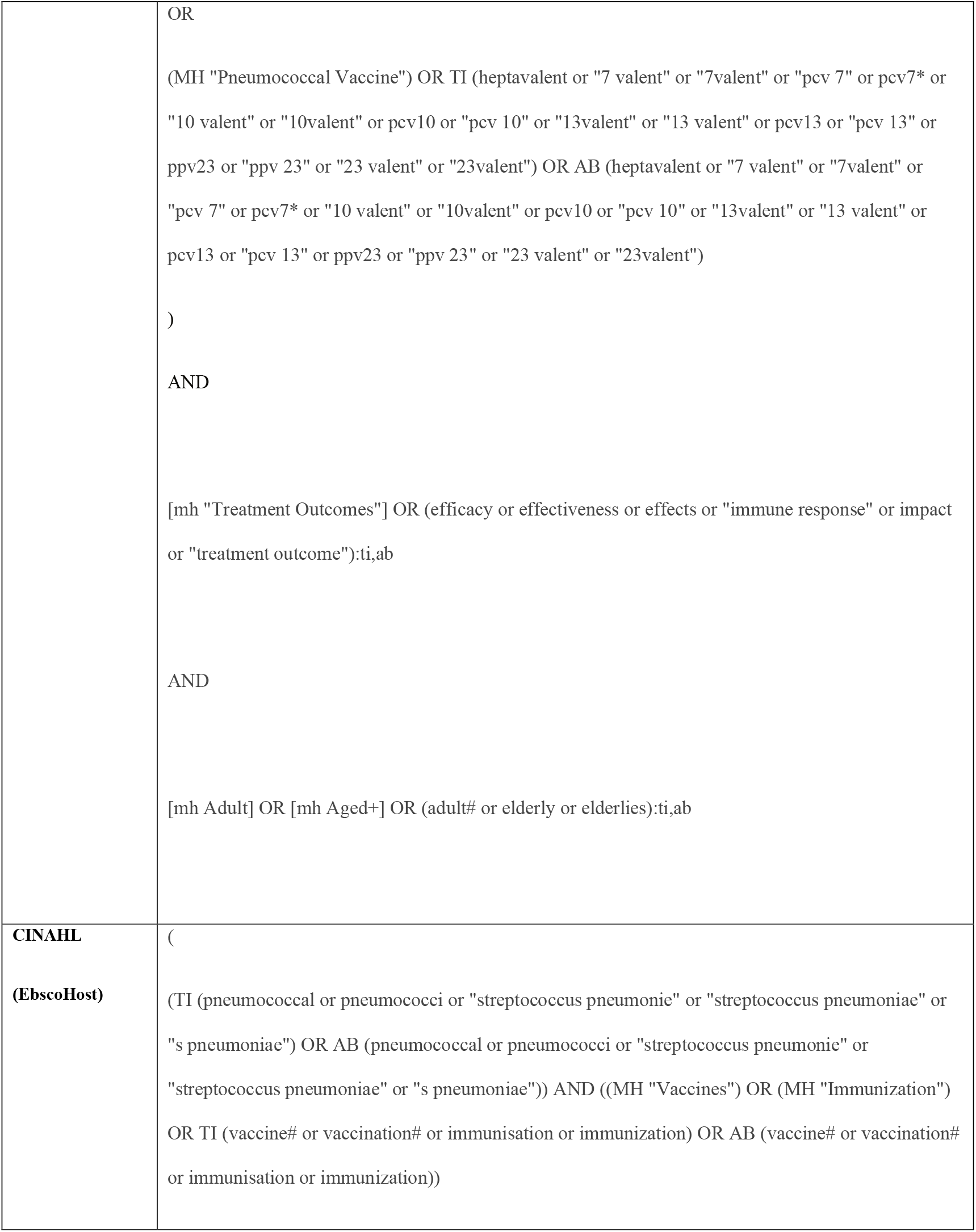
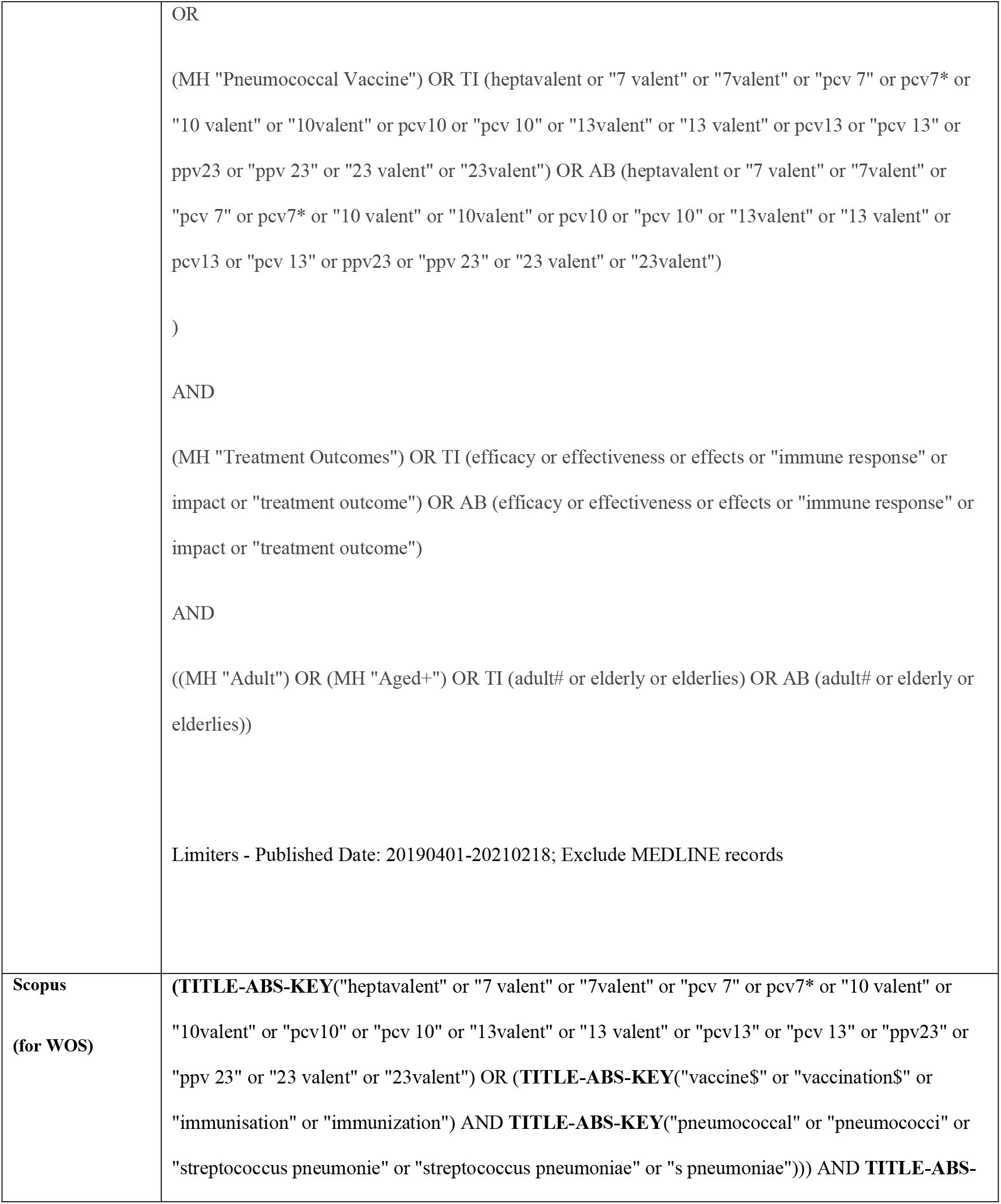
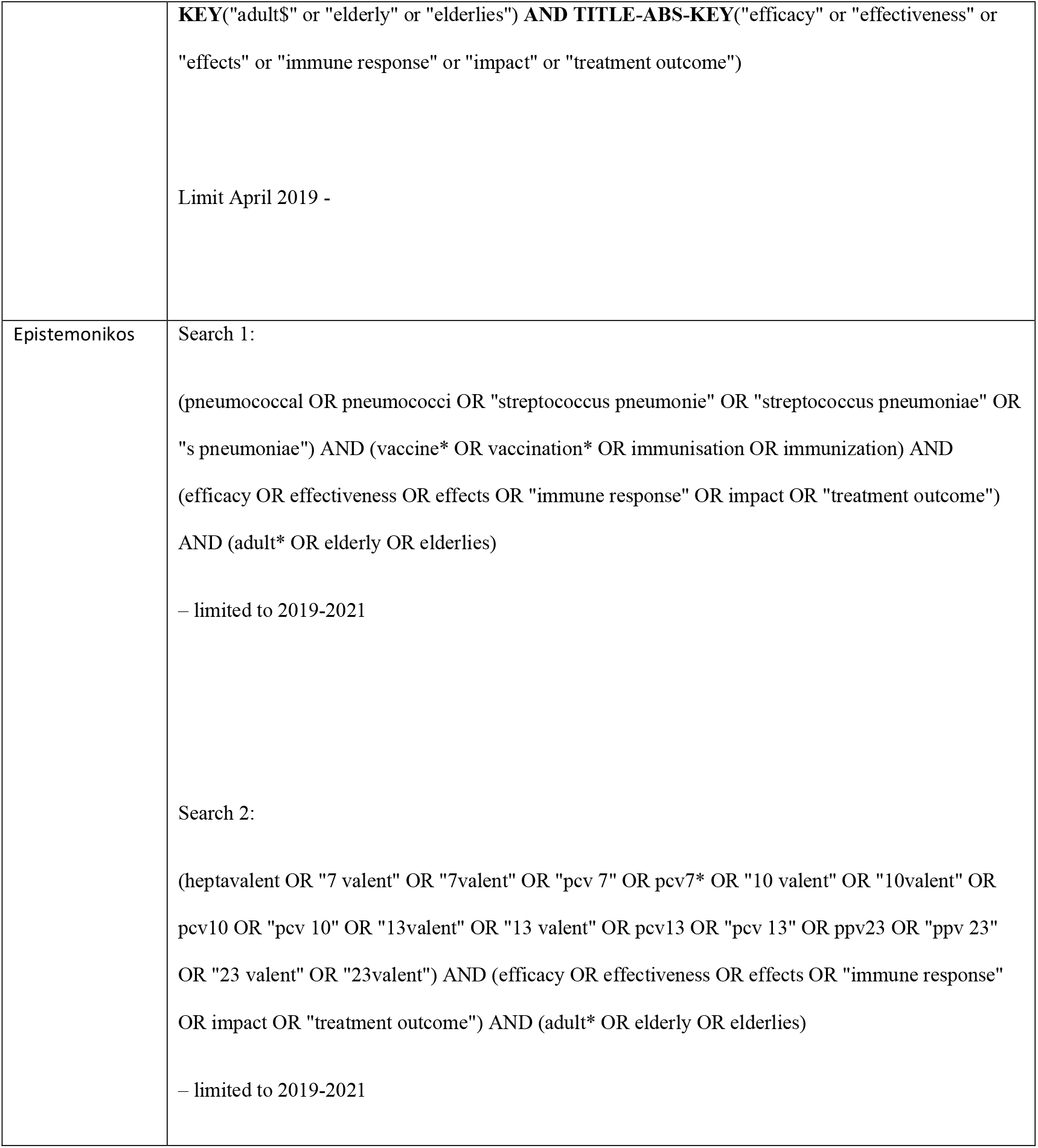

